# The HOSPITAL score and LACE index for predicting the risk of hospital readmission do not show sex discrimination in a retrospective study

**DOI:** 10.1101/2020.02.22.20026740

**Authors:** Robert Robinson

## Abstract

**Introduction:** The perceived absence of human implicit and explicit biases, scalability, and the potential for rapid improvement with algorithmic decision-making systems make compelling arguments for the widespread use of this technology. Unfortunately, real-world performance of some algorithmic decision-making systems demonstrates the reinforcement of discriminatory human biases in a way that is hidden from the human user. This study aims to retrospectively investigate if the widely used HOSPITAL score and LACE index used to predict hospital readmissions exhibit bias on the basis of sex.

**Materials and Methods:** All adult medical patients discharged from the SIU-School of Medicine (SIU-SOM) Hospitalist service from Memorial Medical Center from January 1, 2015, to January 1, 2017, were studied retrospectively to determine if patient sex had an influence on the ability of the HOSPTIAL score and LACE index to predict the likelihood of any cause hospital readmission within 30 days. Receiver operating characteristic (ROC) curves were constructed comparing risk prediction tool performance by sex by measuring the area under the curve (AUC).

**Results:** The analysis includes data for the 1781 discharges for 1410 individual patients that met inclusion criteria. Of these discharges, 456 (27%) were readmitted to the same hospital within 30 days. The overall study population was 47% women, had an average age of 63 years and spent an average of 7.9 days in the hospital. Comparison of the performance of the LACE index in women and men showed no differences between AUCs (0.565 and 0.578, p = 0.613) and an ABROCA of 0.013. Sensitivity (67% and 70%), specificity (46% and 46%), PPV (30% and 31%), NPV (80% and 82%) and accuracy (51% and 52%) for the LACE index are very similar for women and men.

Comparison of the performance of the HOSPITAL in women and men showed no differences between AUCs (0.56 and 0.58, p = 0.407) and an ABROCA of 0.008 indicating highly similar performance. Sensitivity (16% and 21%), specificity (96% and 95%), PPV (59% and 57%), NPV (77% and 78%) and accuracy (76% and 76%) for the HOSPITAL score are very similar for women and men.

**Discussion and Conclusions:** The performance of the HOSPITAL and LACE readmission risk prediction tools appears to have equivalent performance when used for women or men in this small, single-center, retrospective study. Further research is needed to explore the potential of bias and discrimination on risk prediction tools used in healthcare.

## Introduction

The perceived absence of human implicit and explicit biases, scalability, and the potential for rapid improvement with algorithmic decision-making systems make compelling arguments for the widespread use of this technology. Unfortunately, real-world performance of some algorithmic decision-making systems demonstrates the reinforcement of discriminatory human biases in a way that is hidden from the human user.

The most well-publicized example of algorithmic discrimination can be found with the Correctional Offender Management Profiling for Alternative Sanctions (COMPAS) criminal sentencing decision support tool that overestimates the risk of racial minority offenders for committing another crime. These inaccurate predictions have led to higher chances of incarceration and longer sentences for minority offenders in jurisdictions that utilize this software. COMPAS explicitly uses race as a risk of negative outcomes in the criminal justice system, reflecting the real-world data used to develop this decision support system (Angwin et al.; 2016; Garber, 2016; Garcia, 2016; Flores, Bechtel and Lowenkamp 2016; Liptak, 2017).

Algorithmic discrimination on the basis of explicitly stated or deduced minority race or female sex results has been reported with internet search, online shopping, credit applications, job search, employment, and facial recognition (Datta et al. 2018; Chen, Ma, Hannak, Wilson 2018; Lanbrecht and Tucker 2018; Knight 2019). This invisible algorithmic discrimination invisibly reinforces current inequities and limits opportunities for the majority of individuals that interact with these systems.

Decisions in healthcare carry considerable weight and may literally make the difference between life and death for individuals. Improving the effectiveness, equity, and efficiency of healthcare delivery are some of the lofty goals of the digital transformation of healthcare. It seems unlikely that healthcare algorithms would be immune to the biases found in other, less life-critical tasks.

One health care specific algorithm has been evaluated for discrimination in peer-reviewed literature. This unidentified commercial health risk assessment algorithm designed to identify patients who would benefit from additional interventions to improve health was studied extensively by Obermeyer and colleagues. The risk algorithm dramatically overestimated the health of Black patients when compared to White patients by the number of diagnosed illnesses, physiologic parameters, and laboratory data. Negating the race parameter for this algorithm through complex manipulation of data inputs resulted in a 250% increase in the identification of Black patients that would likely benefit from additional healthcare interventions that were known to have positive impacts on individual health (Obermeyer, Powers, Vogeli and Mullainathan 2019).

This study aims to retrospectively investigate if the widely used HOSPITAL score and LACE index used to predict hospital readmissions exhibit bias on the basis of sex. These algorithms have been validated multiple times in studies ranging from a few hundred to over a million patients in patient care settings worldwide (Donzé, Aujesky, William and Schnipper, 2013; Donzé et al., 2016; van Walraven et al., 2010). Nether the HOSPITAL score or LACE index include sex as a predictor variable.

## Materials and Methods

All adult medical patients discharged from the SIU-School of Medicine (SIU-SOM) Hospitalist service from Memorial Medical Center from January 1, 2015, to January 1, 2017, were studied retrospectively to determine if patient sex had an influence on the ability of the HOSPTIAL score and LACE index to predict the likelihood of any cause (avoidable and unavoidable) hospital readmission within 30 days. Exclusion criteria were transferred to another acute care hospital, leaving the hospital against medical advice, or death. Any cause readmission within 30 days of hospital discharge endpoint was selected because it is the measure used by the Medicare HRRP.

Memorial Medical Center is a 507 bed not-for-profit university-affiliated tertiary care center located in Springfield, Illinois, USA. The SIU-SOM Hospitalist service is the general internal medicine residency teaching service staffed by board-certified or board-eligible hospitalist faculty. Patients for the hospitalist service are primarily admitted via the hospital emergency department or transferred from other regional hospitals with acute medical issues. Elective hospital admissions are extremely rare for this service.

Data on age, sex, International Classification of Disease (ICD) diagnosis codes, emergency department visits in the last six months, length of stay, hospital readmission within 30 days and the other variables in the HOSPITAL score (Table 1) and LACE index (Table 2) were extracted from the electronic health record in a de-identified manner for analysis.

**Table 1.** HOSPITAL Score

| Attribute | Points if Positive |
| --- | --- |
| Low hemoglobin at discharge (<12 g/dL) | 1 |
| Discharge from an Oncology service | 2 |
| Low sodium level at discharge (<135 mEq/L) | 1 |
| Procedure during hospital stay (ICD10 Coded) | 1 |
| Index admission type urgent or emergent | 1 |
| Number of hospital admissions during the previous year |  |
| 0-1 | 0 |
| 2-5 | 2 |
| >5 | 5 |
| Length of stay ≥ 5 days | 2 |

**Table 2.** LACE index

| Attribute | Points if Positive |
| --- | --- |
| **Length of stay** |  |
| Less than 1 day | 0 |
| 1 day | 1 |
| 2 days | 2 |
| 3 days | 3 |
| 4-6 days | 4 |
| 7-13 days | 5 |
| ≥ 14 days | 7 |
| **Acute or emergent admission** | 3 |
| **Charlson comorbidity index score** |  |
| 0 | 0 |
| 1 | 1 |
| 2 | 2 |
| 3 | 3 |
| ≥4 | 5 |
| **Visits to the emergency department in the previous 6 months** |  |
| 0 | 0 |
| 1 | 1 |
| 2 | 2 |
| 3 | 3 |
| ≥4 | 4 |

The study hospital does not have a distinct oncology admitting service. To address the increased risk of readmission in oncology patients found in other studies using the HOSPITAL score, this study classified patients with oncology-related diagnosis ICD codes to have been discharged from an oncology service. This reflects local practice patterns where hospitalists often admit patients to the general medicine service for oncologists. Because data is only available from the study hospital, readmissions at other hospitals will not be detected.

Institutional review board review for this study was obtained from the Springfield Committee for Research Involving Human Subjects. This study was determined not to meet the criteria for research involving human subjects according to 45 CFR 46.101 and 45 CFR 46.102.

### Statistical analysis

The influence of sex on the ability of the HOSPITAL score and LACE index to accurately predict any cause hospital readmission within 30 days was investigated. Qualitative variables were compared using Pearson chi^2^ or Fisher’s exact test and reported as frequency (%). Quantitative variables were compared using the nonparametric Mann–Whitney U test and reported as mean ± standard deviation.

The HOSPITAL score and LACE index were calculated for each admission. Readmission risk probability was then assigned based on the outcomes of these calculations.

Receiver operating characteristic (ROC) was constructed comparing risk prediction tool performance by sex by measuring the area under the curve (AUC). The AUC curves were compared for statistically significant differences using the method outlined by DeLong and colleagues (DeLong, Delong, and Clarke-Pearson, 1988).

Absolute Between-ROC Area (ABROCA) values were calculated using the method described by Gardner and colleagues (Gardner, Brooks, and Baker, 2019)

Statistical analyses were performed using R version 3.6.1 (R Foundation for Statistical Computing, Vienna, Austria).

Two sided *P*-values < 0.05 were considered significant.

## Results

During the study period (2 years), 1916 discharges were recorded for the SIU-SOM Hospitalist service. The analysis includes data for the 1781 discharges for 1410 individual patients that met inclusion criteria (Figure 1). Of these discharges, 456 (27%) were readmitted to the same hospital within 30 days. The overall study population was 47% women, had an average age of 63 years and spent an average of 7.9 days in the hospital.

**Figure 1.**
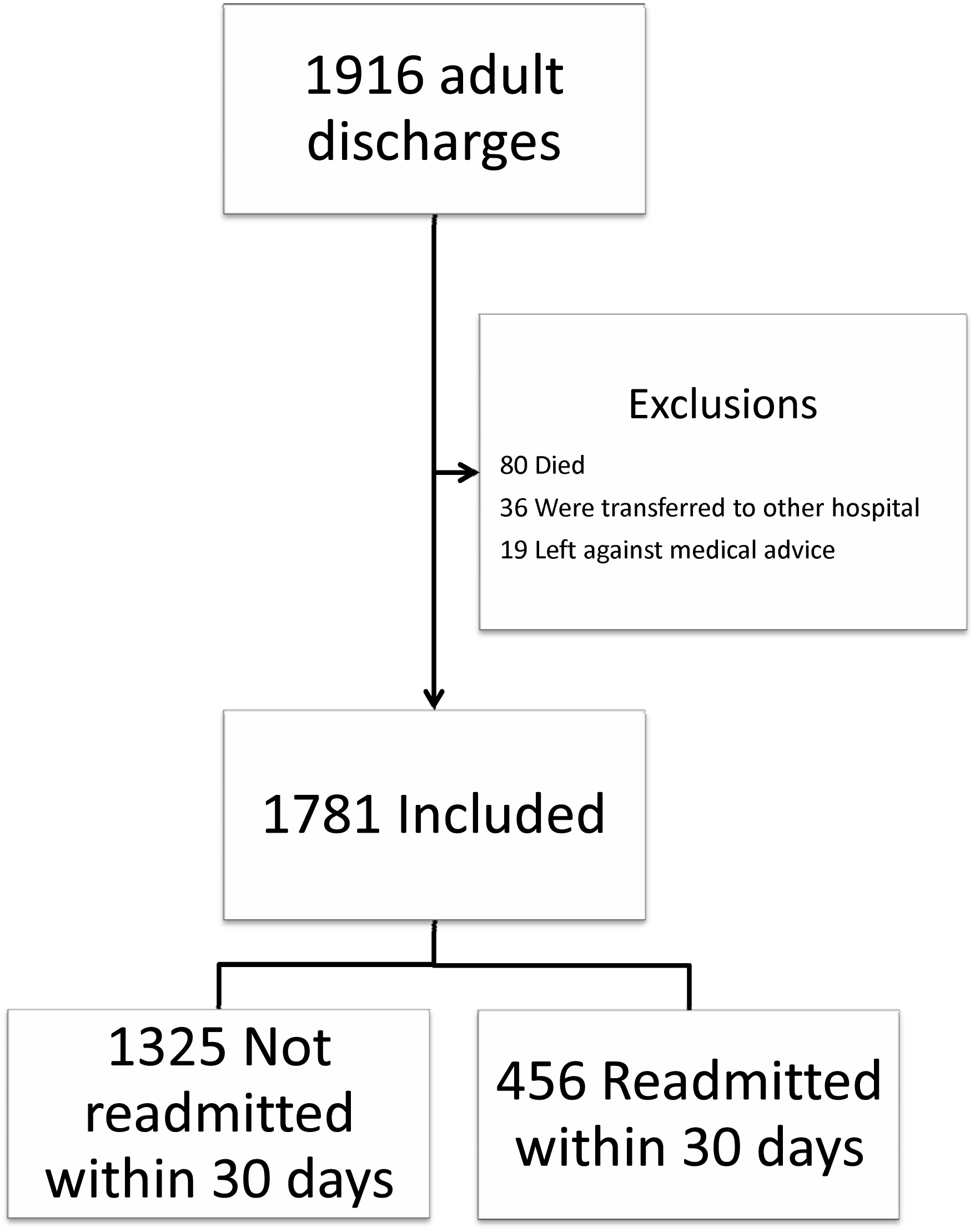
Study Flow Diagram.

The patients readmitted within 30 days of discharge were more frequently admitted to the hospital in the last year, evaluated in the emergency department in the last six months, had higher HOSPITAL scores, higher LACE index values, and higher Charlson Comorbidity Scores. The medical comorbidities of a history of myocardial infarction, CHF, cirrhosis, diabetes, and renal disease were seen more frequently in the patients readmitted within 30 days of hospital discharge. These differences were statistically significant (Table 3).

**Table 3.** Baseline characteristics of the study population by 30-day readmission status

| Characteristic | Not readmitted<br>within 30 days<br>n = 1325 | Readmitted<br>within 30 days<br>n = 456 | p-value |
| --- | --- | --- | --- |
| Age, mean (SD) | 63 (16.0) | 64 (15.5) | 0.144 |
| Women | 624 (47%) | 216 (47%) | 0.919 |
| Length of stay (SD) | 7.7 (7.1) | 8.4 (8.8) | 0.091 |
| Hospital admissions in the last year | 0.71 (0.8) | 1.61 (1.7) | <0.001 |
| Emergency department visits in the last 6 months | 0.39 (1.22) | 1.21 (3.0) | <0.001 |
| HOSPITAL Score | 3.86 (1.4) | 4.96 (1.8) | <0.001 |
| LACE Index | 11.31 (2.4) | 12.59 (3.72) | <0.001 |
| Charlson Comorbidity Score | 4.88 (3.27) | 6 (3.78) | <0.001 |
| Number of discharge medications | 10.33 (6.49) | 11.89 (6.53) | <0.001 |
| Medical Comorbidities |  |  |  |
| Myocardial infarction | 344 (26%) | 150 (33%) | 0.004 |
| Congestive heart failure | 295 (22%) | 159 (35%) | <0.001 |
| Peripheral artery disease | 118 (9%) | 45 (10%) | 0.539 |
| Stroke | 76 (6%) | 26 (6%) | 0.978 |
| Dementia | 40 (3%) | 9 (2%) | 0.239 |
| Chronic lung disease | 357 (27%) | 144 (32%) | 0.058 |
| Connective tissue disease | 28 (2%) | 7 (2%) | 0.443 |
| Peptic ulcer disease | 59 (5%) | 19 (4%) | 0.797 |
| Cirrhosis | 40 (3%) | 25 (6%) | 0.016 |
| Diabetes without complications | 272 (21%) | 127 (28%) | 0.001 |
| Diabetes with complications | 145 (11%) | 88 (19%) | <0.001 |
| Paralysis | 44 (3%) | 24 (5%) | 0.062 |
| Renal disease | 243 (18%) | 142 (31%) | <0.001 |
| Cancer | 97 (7%) | 42 (9%) | 0.194 |
| Metastatic cancer | 32 (2%) | 18 (4%) | 0.088 |

Comparison of the performance of the LACE index in women and men showed no differences between AUCs (0.565 and 0.578, p = 0.613, Figure 2) and an ABROCA of 0.013 indicating highly similar performance over the full range of LACE index values (Table 4). Sensitivity (67% and 70%), specificity (46% and 46%), PPV (30% and 31%), NPV (80% and 82%) and accuracy (51% and 52%) for the LACE index are very similar for women and men.

**Table 4.** LACE index performance by sex

|  | Women | Men |
| --- | --- | --- |
| **Sensitivity** | 67% | 70% |
| **Specificity** | 46% | 46% |
| **Positive Predictive Value** | 30% | 31% |
| **Negative Predictive Value** | 80% | 82% |
| **Accuracy** | 51% | 52% |

**Figure 2.**
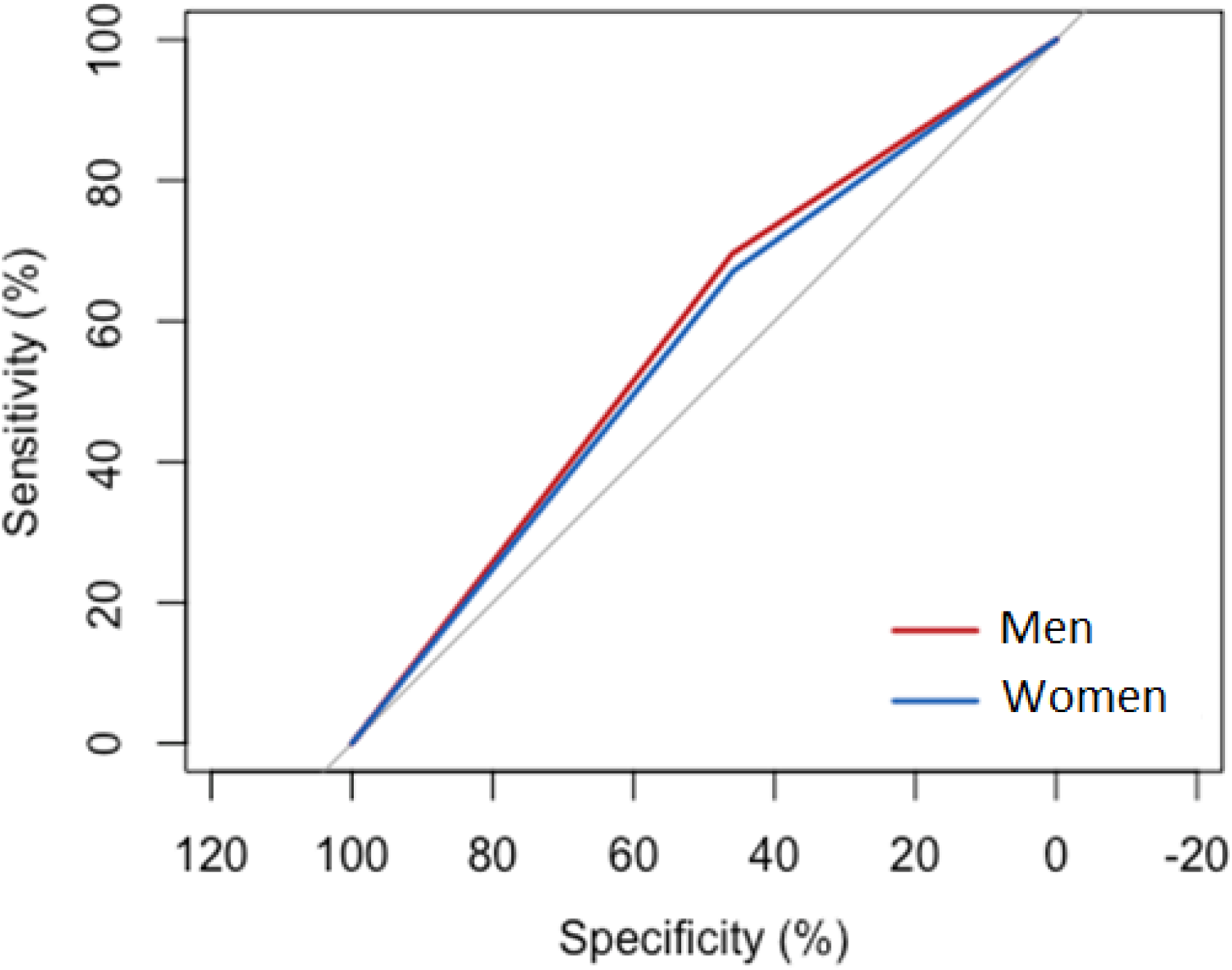
Receiver operating characteristic curve for the LACE index by sex.

Comparison of the performance of the HOSPITAL in women and men showed no differences between AUCs (0.56 and 0.58, p = 0.407, Figure 3) and an ABROCA of 0.008 indicating highly similar performance over the full range of HOSPITAL score values (Table 5). Sensitivity (16% and 21%), specificity (96% and 95%), PPV (59% and 57%), NPV (77% and 78%) and accuracy (76% and 76%) for the HOSPITAL score are very similar for women and men.

**Table 5.** HOSPITAL Score performance by sex

|  | Women | Men |
| --- | --- | --- |
| **Sensitivity** | 16% | 21% |
| **Specificity** | 96% | 95% |
| **Positive Predictive Value** | 59% | 57% |
| **Negative Predictive Value** | 77% | 78% |
| **Accuracy** | 76% | 76% |

**Figure 3.**
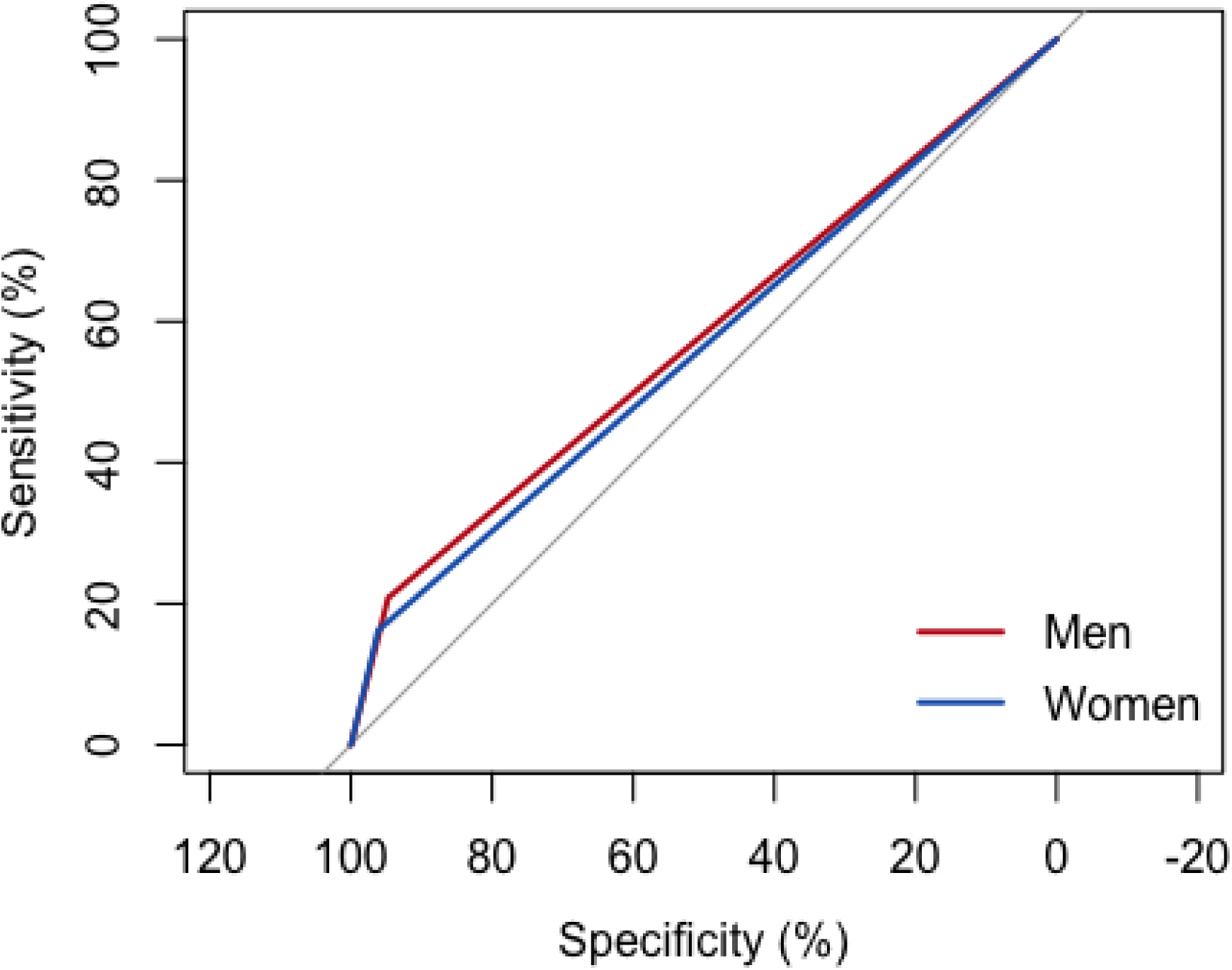
Receiver operating characteristic curve for the HOSPITAL score by sex.

## Discussion

This study has several important limitations. This study and the international validation study for the HOSPITAL score share an important shortfall by only identifying readmissions within 30 days at the same hospital (Donzé et al., 2016). Furthermore, this study is retrospective, single-center, focused on medical patients, small sample size, and shaped by local practice patterns (no oncology admitting service, few elective admissions). These limitations may reduce the generalizability of these results.

This study shows the performance of the HOSPITAL and LACE readmission risk assessment tools do not demonstrate sex discrimination when predicting the risk of hospital readmission within 30 days of discharge.

## Conclusions

The performance of the HOSPITAL and LACE readmission risk prediction tools appears to have equivalent performance when used for women or men in this small, single-center, retrospective study.

Further research is needed to explore the potential of bias and discrimination on risk prediction tools used in healthcare.

## Data Availability

Data is available for review on request to the authors.

